# The Advanced Organ Support (ADVOS) hemodialysis system fulfills its intended purpose: Analysis of data from 282 patients from the Registry on Extracorporeal Multiple Organ Support (EMOS)

**DOI:** 10.1101/2025.01.26.25321153

**Authors:** Valentin Fuhrmann, Bartosz Tyczynski, Aritz Perez Ruiz de Garibay, Tobias Michael Bingold, Julia Weinmann-Menke, Andreas Faltlhauser, Dominik Jarczak, Jens Lutz, Michael Sander, Pascal Klimpke, Christian Koch, Andreas Kribben, Olaf Boenisch, Stefan Kluge

## Abstract

**Background:** Several case series have highlighted the ADVOS hemodialysis system’s efficacy in eliminating water-soluble and protein-bound substances across diverse patient populations, such as multiorgan failure, acute-on-chronic liver failure (ACLF), acidosis, and even COVID-19. The EMOS-Registry, a non-interventional, multi-center patient registry, amassed real-world evidence, culminating in the largest patient cohort treated with ADVOS to date. This study aims to present and analyze the final performance and safety outcomes from the entire dataset comprising 282 participants.

**Methods:** Data spanning from January 18, 2017, to August 31, 2020, were collected from five German hospitals, encompassing subsets of patients with acidosis and ACLF grade 3. Performance and safety were assessed through vital signs, clinical laboratory parameters and blood gas analyses. The SOFA Score-Standardized Mortality Ratio (SMR) served to evaluate patient outcomes in the absence of a control group.

**Results:** Participants, with a median age of 58 years, predominantly male (64%), exhibited a high requirement for mechanical ventilation (68%) and vasopressors (82%) with a median SOFA Score of 15. Notably, a median of 3 (IQR 2, 5) ADVOS sessions per patient were administered.

Following the initial treatment, significant reductions were observed in bilirubin (–1.9 [CI 95% –1.3, – 2.5]), creatinine (–0.5 [-0.4, –0.6]), and blood urea nitrogen (–13.1 mg/dL [-10.3, –16.0]) levels. Moreover, there were marked improvements in blood pH (7.34 vs. 7.41, p<0.001), HCO3-(19.4 vs. 24.6 mmol/l, p<0.001) and base excess (–5.6 vs. 0.2 mmol/l, p<0.001). The observed mortality rate (66%) was notably lower than the expected rate based on SOFA Score (84%), resulting in a SMR of 0.79 (95% CI: 0.66-0.93), with a calculated number needed to treat (NNT) of 5.8.

**Conclusions:** This study emphasizes the ADVOS system’s efficacy in eliminating water-soluble and protein-bound substances and correcting acid-base imbalances across a diverse cohort with multiorgan failure. However, further validation through randomized controlled trials is warranted to solidify these findings.

*Trial registration: DRKS00017068. Registered 29 April 2019 – Retrospectively registered,* https://drks.de/search/en/trial/DRKS00017068

## 1. Background

The ADVOS multi hemodialysis system (ADVITOS GmbH, Munich, Germany) effectively combines kidney, liver, and lung support together with acid-base balance correction for patients with multiple organ failure within a single device [1]. In this extracorporeal organ support procedure, a circulating and reusable electrolyte solution enriched with albumin functions as dialysate fluid. Its purpose is to eliminate protein-bound toxins from the bloodstream [2]. This distinctive approach differs from traditional dialysis methods, as it not only targets water-soluble substances (such as creatinine, urea, and ammonia), but also addresses albumin-bound compounds (such as bilirubin, bile acids, aromatic amino acids, and copper) [3–5]. Furthermore, an innovative dialysate recirculation and recycling circuit enables the customization of the dialysate’s pH value and composition (i.e., bicarbonate content) for individual patients. This capability facilitates the management of acid-base balance, including the correction of metabolic acidosis and the fluid-based removal of CO_2_ [6].

In the last decade, several case-series have reported the benefits of ADVOS regarding removal of water-soluble and protein-bound substances in different populations, including patients with multiorgan failure, acute-on-chronic liver failure, acidosis or even COVID-19 [7–10]. Data are collected from retrospective studies with small sample sizes aimed to assess feasibility and safety. In addition to these, a non-interventional, multi-center, and non-randomized patient registry gathered real-world evidence in patients with an indication for multiple organ dialysis with the ADVOS device. The preliminary two year analysis reported beyond feasibility and safety, a trend towards mortality rate reduction in patients treated with ADVOS [11]. It was concluded that due to the nature of patient’s registries, the data should be carefully interpreted.

In the absence of prospective data from randomized controlled trials, patient registries offer a valuable alternative or complement for data collection as they can provide insights into real-world treatment outcomes from broader patient populations [12]. This is also applicable to patients treated with blood purification techniques [13–17]. Moreover, they can help to generate hypotheses and guide investigators to identify factors that may warrant more focused research.

The use of risk-adjustment methods such as the standardized mortality ratio (SMR) can be valuable tools for the analysis of patient-centered outcomes when a control group is lacking [18]. Given the heterogenicity of real-world data, risk-adjusted mortality rates help to account for the influence of various patient characteristics and comorbidities. This ensures that observed mortality rates are more closely aligned with the true effectiveness of the intervention rather than being distorted by differences in patient health status. In any case, the risk-adjustment method should be carefully chosen and appropriately validated for the specific patient population [19]. In this regard, the Sequential Organ Failure Assessment (SOFA) Score is a widely recognized and validated tool used to assess the severity of organ dysfunction in critically ill patients [20]. It evaluates the functioning of several organ systems, including the respiratory, cardiovascular, hepatic, coagulation, renal, and neurological systems [21]. The SOFA Score is commonly used to monitor the progression of organ failure and to predict patient outcomes, including mortality [22, 23].

The objective of this work is to present and discuss the final performance and safety data from the Extracorporeal Multiple Organ Support (EMOS) patient registry with the ADVOS hemodialysis system. Special focus is made in the analysis of risk-adjusted mortality considering a SOFA Score-standardized mortality ratio. In addition, data from subgroups of patients with acute-on-chronic liver failure (ACLF) or acidosis (two of the main core groups of patients treated with ADVOS) are described.

## 2. Methods

### 2.1 Study design, setting and participants

The current analysis considered the data from the whole patient cohort from the EMOS-Registry collected between January 18, 2017, and August 31, 2020, in five German hospitals. It included subsets of data from patients with acidosis (i.e., blood pH < 7.35) and patients with ACLF grade 3. The EMOS registry was a non-interventional, multi-center, non-randomized registry in post marketing surveillance aimed to collect data on real-life treatment conditions for patients for whom multiple organ dialysis with the ADVOS hemodialysis system was indicated.

The purpose of the EMOS-registry was to assess the efficacy and safety of the procedure using real-world data. Additionally, the registry aimed to formulate recommendations for ADVOS treatments and identify suitable supportive and diagnostic strategies. Moreover, the evaluation sought to compare the registry’s real-life practice data on adverse events, mortality rates, and treatment outcomes with the findings from other published trials.

The methodological details of the EMOS-Registry were recently reported [11]. Briefly, no exclusion criteria was defined and all adult patients (i.e. ≥ 18 years) with an indication for the ADVOS hemodialysis system and at discretion of the treating physician, were enrolled in the registry. The intended purpose of the ADVOS hemodialysis system comprises the removal of water-soluble toxic substances, protein-bound toxic substances, the normalization or improvement of the composition of blood in case of e.g., electrolyte or acid-base disturbances (e.g., metabolic acidosis or respiratory acidosis) and the removal of fluids in case of fluid overload. It is intended to be used in healthcare facilities in patients with acute, chronic and acute-on-chronic liver failure and/or renal failure and/or acidosis. It is not intended for use in children, or patients with blood volume ≤ 4.3 liter, pregnant women, nursing woman and patients with prion disease (e. g. Creutzfeldt-Jakob disease).

Eligible patients were connected to the blood tubing set of the ADVOS multi through a conventional double lumen dialysis catheter (e.g., 13 F diameter). Blood flows between 100 and 400 mL/min through two parallel high-flux dialyzers with a 1.9 m^2^ effective surface each, either continuously up to 24 hours or during intermittent treatment sessions. Anticoagulation is strongly recommended during treatments and was employed on clinical judgement.

### 2.2 Variables and measurements

#### Baseline characteristics

The participants were defined using data on age, gender, prognostic health scores (e.g., SOFA), required interventions (e.g., need for vasopressors and mechanical ventilation), the presence of acidosis or acute-on-chronic liver failure at baseline and documented comorbidities at hospital admission, among others.

#### Performance

The study did not have any pre-defined primary or secondary endpoint, as its main objective was to gather real-life data on multiple organ dialysis. Performance and safety were evaluated using data on vital signs, clinical laboratory variables (e.g., bilirubin, creatinine, blood urea nitrogen) and blood gas analyses (e.g., pH, serum bicarbonate, pCO2, base excess). The complete assessment table can be found in the already published 2-year analysis [11]. After each patient’s final ADVOS treatment session, the specific settings used (e.g., blood flow, dialysate pH, treatment duration) were summarized and reported.

#### Safety

The reporting of Adverse Events (AEs) was limited to catheter problems, bleeding, allergic reactions, clotting, electrolyte imbalances, and infections for all the patients exposed to the registry medical device.

#### SOFA Score-standardized mortality ratio (SOFA-SMR)

In the absence of a control group, it was deemed appropriate to evaluate the benefits of the treatment with the ADVOS hemodialysis system through the standardized mortality ratio based on the SOFA Score at baseline (i.e., immediately before the first ADVOS session).

The Standardized Mortality Ratio is a statistical measure used to compare the mortality rates of a specific population with that of a standard or reference population. It is a common method to assess and understand the relative risk of death in different groups or areas.

To calculate the SMR, the observed number of deaths in the treated patients was compared to the expected number of deaths that would have occurred considering their SOFA score at baseline (i.e., immediately before the first ADVOS session). For calculation purposes, this SOFA Score considered as being the highest. Patients were divided into groups based on their expected mortality rate as shown by Ferreira et al [23]. The expected number of events (i.e., deaths) was calculated in each group and then added to obtain the total number of expected events.

The formula for calculating the Standardized Mortality Ratio (SMR) is:

SMR = (Observed number of deaths / Expected number of deaths)

If the SMR is equal to 1, it indicates that the observed mortality is precisely what would be expected according to the SOFA Score. A value greater than 1 suggests that the treated patients had a higher mortality rate than the standard population, while a value less than 1 indicates a lower mortality rate.

Moreover, the Absolute Risk Reduction (ARR) and Number Needed to Treat (NNT) were calculated based on data from expected and observed deaths. ARR represents the actual reduction in the risk of death attributable to a specific treatment or intervention. When positive, it means that the treatment or intervention reduces the risk of the outcome (death) compared to that expected. It was calculated as follows:

ARR = Expected deaths – Observed deaths

The NNT is a measure that quantifies how many patients need to receive a specific treatment to prevent one adverse outcome (e.g., death) over a defined period. It gives an idea of the practical impact of a treatment by indicating how many patients might benefit from the intervention. A low NNT suggests that the treatment is effective in preventing adverse outcomes, as few patients need to be treated to prevent one event. The following formula was employed:

NNT = 1 / ARR

The SMR, the ARR and the NNT were calculated only for those patients with documented SOFA scores at baseline.

### 2.3 Study population and subgroups

According to the study plan, at least 100 patients from 3 centers were expected to be enrolled during a 2-year period, with the possibility of a yearly extension after data review [11]. As of August 31, 2020, a total of 282 patients were enrolled in the study, with participation from 5 clinical sites in Germany.

Specifically, the University Hospital Hamburg-Eppendorf (UKE) enrolled 169 patients, the Mainz University Medical Center from the Johannes Gutenberg University included 35 patients, the University Hospital in Essen registered 20 patients, the Weiden Clinic (Kliniken Nordoberpfalz AG) contributed with 30 patients and the University Clinic in Gießen documented data from 28 patients.

Different subgroup analyses were performed based on the indications of the ADVOS hemodialysis system. This includes patients with kidney failure, liver failure and acidosis. The overall evaluation comprised 282 patients with an indication for hemodialysis with the ADVOS system and were considered to have kidney injury. Since the exact diagnosis was not documented in the registry, patients with a preexisting mild, moderate or severe liver disease [24] at hospital admission and an ACLF Grade 3 according to the CLIF Organ Failure score at baseline were included in the ACLF 3 group (n = 53) [25]. Additional subgroups of patients with ACLF Grade 2 (n=26) or Grade 1 (n=8) were not specifically analyzed due to the small number of subjects included. Finally, the subgroup of patients with acidosis included participants with pH < 7.35 at baseline (n = 146).

### 2.4 Statistical analysis

Continuous variables are presented as median and interquartile range (IQR). The normal distribution of samples was assessed using the Shapiro–Wilk test, while the Levene test was utilized to evaluate the homogeneity of variance. To compare values before and after ADVOS sessions and treatments, the Student’s t-test for paired samples was employed. Variables that did not show homogeneous distribution were compared using the Mann-Whitney-U-Test. A two-tailed p-value below 0.05 was considered statistically significant. 28-day and overall mortality is shown as percentage and as SMR and 95% confidence intervals (CI 95%). Data were analyzed with IBM SPSS 28.0 for Windows®.

### 2.5 Ethical principles, patient safety, data protection and funding

This study received approval from the Bavarian State Medical Association (Bayerische Landesärztekammer) on November 2, 2016, and is registered in the German Registry for Clinical Studies and the International Clinical Trials Registry Platform of the World Health Organization (DRKS00017068). The registry does not entail any additional risks to the patients beyond those associated with data collection and storage. Patients were treated in accordance with approved guidelines, and their treating physicians had already determined that treatment with multiple organ dialysis was warranted.

All participating patients who were scheduled for pseudonymous data collection were required to provide informed consent by signing a consent form. For those without consent, no personal data related to the registry can be accessed due to the anonymized nature of the data.

Patient confidentiality was strictly adhered to in accordance with the European Data Protection Directive and other relevant international and national requirements [26]. During the course of this registry, necessary amendments were made to comply with the new EU General Data Protection Regulation [27].

The study was sponsored by ADVITOS GmbH (formerly Hepa Wash GmbH). This project has received funding from the European Union’s Horizon 2020 research and innovation programme under grant agreement No 880349.

## 3. Results

### 3.1 Baseline characteristics

The median age of participants in the registry was 58 years (IQR 48, 68) and 64% were male. Patients were mainly admitted after being transferred from another hospital (35%) and from the emergency ward (45%). The patients were critically ill at baseline. 68% required mechanical ventilation and 82% vasoactive substances. All 282 patients had an indication for dialysis. From them, 52% of the patients had a pH < 7.35 at baseline. Independently of the presence of acidosis, 87 patients had a documented acute-on-chronic liver failure at either Grade 1 (8), Grade 2 (26) or Grade 3 (53). Among the recruited participants, SOFA Score was documented in 202 patients, with a median value of 15 (IQR 12, 18). Additional baseline characteristics as well as medical history at hospital admission are summarized in Table 1 and Supplementary Table 1.

**Table 1.** Baseline characteristics immediately before the 1st ADVOS treatment session in the whole data set (n=282). Median (IQR) or percentage. *Incomplete data at baseline did not allow to calculate the CLIF Organ Failure Score in many cases. This may lead to an underestimation of the number of ACLF patients

| Parameters | Median | IQR 25 | IQR 75 |
| --- | --- | --- | --- |
| Age | 58 | 48 | 67 |
| Sex (male, %) | 181 | 64% |  |
| Reason for admission (n, %) |  |  |  |
| Emergency | 127 | 45% |  |
| Planned admission (e.g. diagnostic, surgery) | 49 | 17% |  |
| Bridging to Transplantation | 7 | 2% |  |
| Transfer from other hospital | 98 | 35% |  |
| Alcohol abuse (n, %) | 114 | 40% |  |
| Smoker (n, %) | 61 | 22% |  |
| Body height | 175 | 167 | 180 |
| Body weight | 80 | 70 | 90 |
| Vasoactive substances (n, %) | 231 | 82% |  |
| Mechanical ventilation (n, %) | 191 | 68% |  |
| Acidosis (n, %) | 146 | 52% |  |
| Acute-on-chronic-liver failure (ACLF)* |  |  |  |
| pre-existing liver disease | 167 | 59% |  |
| CLIF-C-ACLF Score documented | 88 | 31% |  |
| CLF-C-ACLF Score | 61 | 51 | 67 |
| no ACLF | 1 | 0% |  |
| ACLF grade 1 | 8 | 3% |  |
| ACLF grade 2 | 26 | 9% |  |
| ACLF grade 3 | 53 | 19% |  |
| Glasgow Coma Score | 9 | 3 | 15 |
| Hepatic Encephalopathy Grade | 1 | 0 | 1 |
| No Encephalopathy | 121 | 43% |  |
| Grade 1 | 18 | 6% |  |
| Grade 2 | 24 | 9% |  |
| Grade 3 | 16 | 6% |  |
| Grade 4 | 6 | 2% |  |
| SOFA Score | 15 | 12 | 18 |
| SOFA Cardiac | 4 | 3 | 4 |
| SOFA Respiratory | 3 | 2 | 3 |
| SOFA Liver | 3 | 2 | 4 |
| SOFA Kidney | 3 | 1 | 4 |
| SOFA Coagulation | 2 | 0 | 3 |
| SOFA GCS | 3 | 0 | 4 |
| Comorbidities at hospital admission (n, %) |  |  |  |
| Myocardial infarction | 16 | 6% |  |
| Congestive heart failure | 50 | 18% |  |
| Peripheral vascular disease | 20 | 7% |  |
| Cerebrovascular disease | 16 | 6% |  |
| Dementia | 1 | 0,4% |  |
| Chronic pulmonary disease | 36 | 13% |  |
| Rheumatologic disease | 6 | 2% |  |
| Peptic ulcer disease | 30 | 11% |  |
| Mild liver disease | 32 | 11% |  |
| Diabetes without chronic complications (without end organ damage) | 66 | 23% |  |
| Diabetes with chronic complications (with end organ damage) | 8 | 3% |  |
| Moderate or severe renal (kidney) disease | 49 | 17% |  |
| Hemiplegia or paraplegia | 5 | 2% |  |
| Malignancy (during last 5 years) | 39 | 14% |  |
| Leukemia | 4 | 1% |  |
| Lymphoma | 8 | 3% |  |
| Moderate or severe liver disease | 139 | 49% |  |
| Metastatic solid tumor | 12 | 4% |  |
| AIDS / HIV | 2 | 1% |  |

### 3.2 ADVOS Treatment settings and safety

A total of 1075 ADVOS treatment sessions were documented, with each patient receiving a median of 3 (IQR 2 to 5) sessions with a median duration of 19 (IQR 10 to 23) hours per session (Table 2 and Supplementary Table 2). The median average blood flow was 100 mL/min and did not vary among the different subgroups. The median dialysate pH set was 7.8 (IQR 7.4, 8.5) for the overall population and 8.0 (IQR 7.6, 9.0) for patients with acidosis. 84% of the treatment sessions were conducted as planned without abortions. In detail, 6.7% of the treatments were aborted by device related issues, 1.6% due to dialyzer inflow related causes, 1.3% due to problems with the catheter and 5.9 % due to other causes, including clotting issues, emergency interventions or death not related to treatment, among others.

**Table 2.** ADVOS Treatment settings. Median (IQR). The UF rate also includes the volume corresponding to potential glucose, citrate or calcium administration, which account to approximately 70-120 mL/h.

| **Treatment settings** | **Median** | **IQR 25** | **IQR 75** |
| --- | --- | --- | --- |
| Total number of treatment sessions | 1075 |  |  |
| Treatment session/patient | 3 | 2 | 5 |
| Median treatment duration (h) | 19 | 10 | 23 |
| Median blood flow (mL/min) | 100 | 100 | 150 |
| Median concentrate flow (mL/min) | 160 | 160 | 200 |
| Median pH value | 7,8 | 7,4 | 8,5 |
| Median Ultrafiltration rate (mL/h) | 135 | 70 | 255 |
| Average Ultrafiltration volume (mL) | 2500 | 650 | 4882 |
| Patients with treatment abortions (n, %) | 108 | 38% |  |
| Total number of treatment abortions (n, %) | 167 | 16% |  |

During the course of the treatments, a total of 151 adverse events were documented. According to the treating physicians, 26 of these events were device-related and were characterized as clotting problems (25) and a single bleeding episode (Table 3).

**Table 3.** Adverse Events documented during ADVOS sessions. The percentage refers to the total number of sessions.

| **Adverse events** | **n** | **%** |
| --- | --- | --- |
| Number of adverse events | 151 | 4% |
| Catheter problems | 9 |  |
| Bleeding | 21 |  |
| Allergic reaction | 1 |  |
| Clotting | 75 |  |
| Electrolyte imbalance | 29 |  |
| Infection | 16 |  |
| Number of device-related adverse events | 26 | 0,2% |
| Catheter problems | 0 |  |
| Bleeding | 1 |  |
| Allergic reaction | 0 |  |
| Clotting | 25 |  |
| Electrolyte imbalance | 0 |  |
| Infection | 0 |  |

### 3.3 Performance of ADVOS according to its intended use

#### Removal of water-soluble and protein-bound toxic substances

A significant removal of bilirubin, creatinine, and blood urea nitrogen (BUN) was documented during the first ADVOS session (Table 4). The reduction rate was concentration dependent (Supplementary Figure 1). The documented removal was higher in those patients with concentrations above the median values for both bilirubin (8% vs. 18%), creatinine (18% vs, 26%) and BUN (29% vs. 33%). Median values at each timepoint are shown in Table 5 and paired differences for first ADVOS treatments in Table 6. These removal rates resulted in a significant reduction of bilirubin, creatinine, and BUN concentrations with mean differences after the first treatment of –1.9 (CI 95%: –1.3, –2.5), –0.5 (CI 95%: –0.4, –0.6) and –13.1 (CI 95%: –10.3, –16.0) mg/dL, respectively (Table 6). Moreover, in patients with ACLF Grade 3, who had even higher baseline bilirubin concentrations, a higher reduction rate per session and mean difference from baseline to post 1^st^ treatment (–3.0 mg/dL, CI 95%: –1.3, –4.7) was observed (Table 6).

**Table 4.** Reduction rates after the first ADVOS treatment for bilirubin, creatinine, and urea in the whole data set and in each of the subgroups. n = number of available pairs of measurements at baseline and Post 1^st^ treatment.

|  |  | ALL |  |  |  | Acidosis |  |  |  | No Acidosis |  |  |  | ACLF 3 |  |  |  |
| --- | --- | --- | --- | --- | --- | --- | --- | --- | --- | --- | --- | --- | --- | --- | --- | --- | --- |
|  |  | Median | IQR 25 | IQR 75 | n | Median | IQR 25 | IQR 75 | n | Median | IQR 25 | IQR 75 | n | Median | IQR 25 | IQR 75 | n |
| Bilirubin total | Baseline-Post 1st Treatment | -8% | -25% | 13% | 200 | -3% | -23% | 22% | 95 | -13% | -27% | 3% | 105 | -14% | -23% | 3% | 39 |
| Creatinine | Baseline-Post 1st Treatment | -18% | -36% | 0% | 187 | -26% | -44% | -8% | 90 | -14% | -29% | 5% | 97 | -23% | -36% | -6% | 38 |
| BUN | Baseline-Post 1st Treatment | -29% | -46% | -10% | 147 | -32% | -61% | -16% | 73 | -25% | -38% | -3% | 74 | -29% | -44% | -13% | 34 |

**Table 5.**
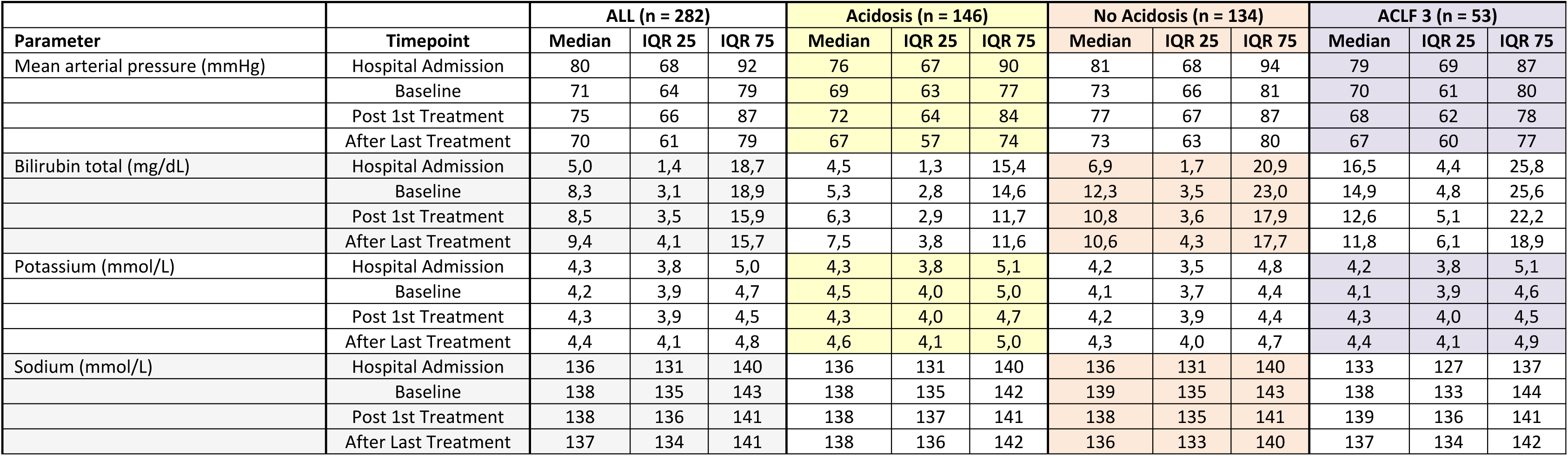
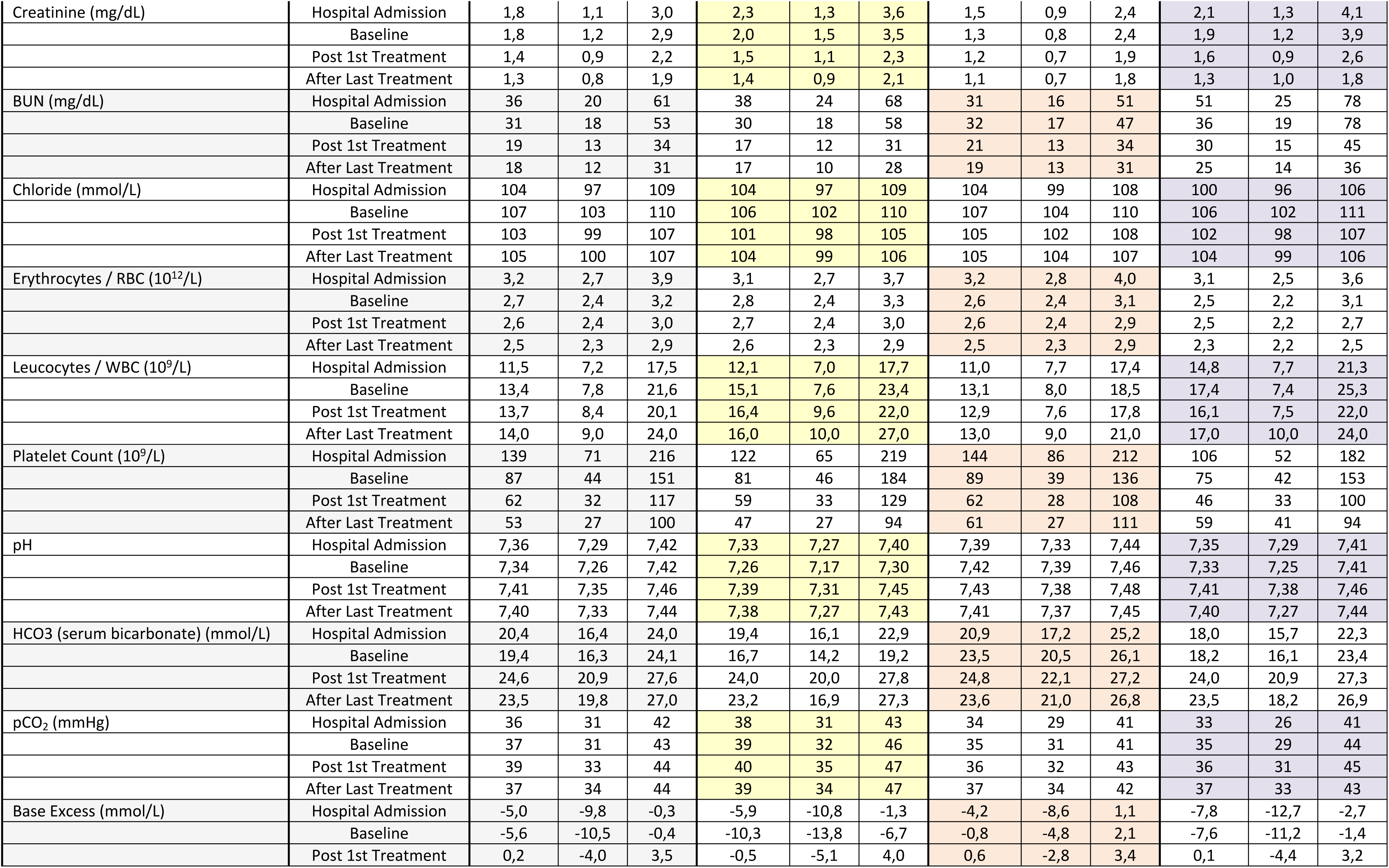
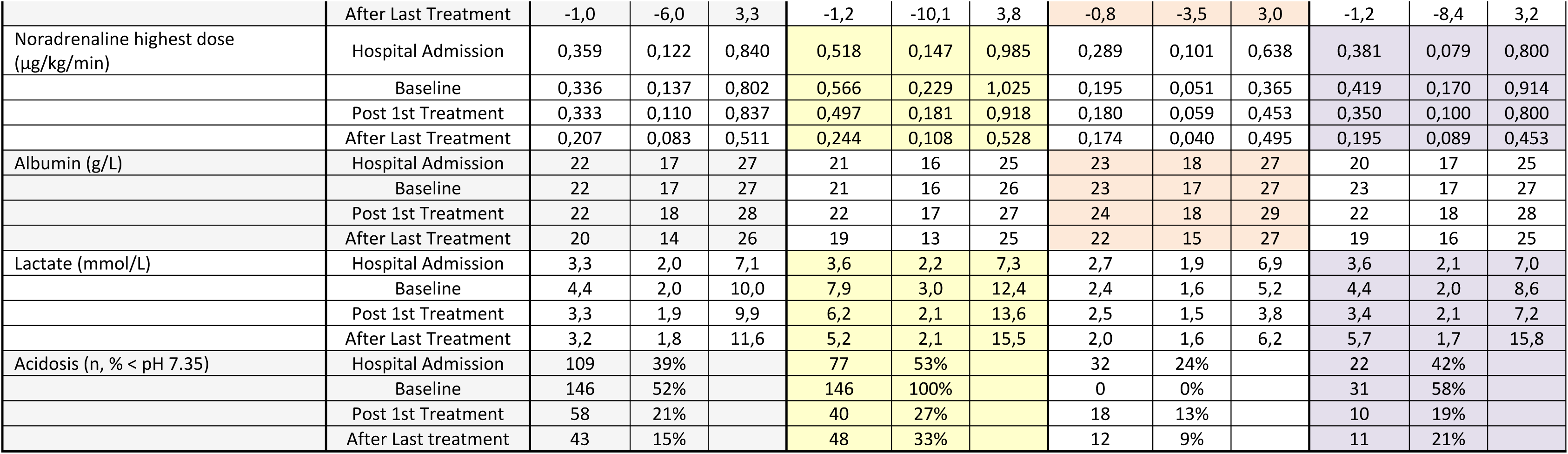
Course of treatment performance parameters in the whole data set and in each of the subgroups. Median (IQR).

| Parameter | Timepoint | ALL (n = 282) |  |  | Acidosis (n = 146) |  |  | No Acidosis (n = 134) |  |  | ACLF 3 (n = 53) |  |  |
| --- | --- | --- | --- | --- | --- | --- | --- | --- | --- | --- | --- | --- | --- |
|  |  | Median | IQR 25 | IQR 75 | Median | IQR 25 | IQR 75 | Median | IQR 25 | IQR 75 | Median | IQR 25 | IQR 75 |
| Mean arterial pressure (mmHg) | Hospital Admission | 80 | 68 | 92 | 76 | 67 | 90 | 81 | 68 | 94 | 79 | 69 | 87 |
|  | Baseline | 71 | 64 | 79 | 69 | 63 | 77 | 73 | 66 | 81 | 70 | 61 | 80 |
|  | Post 1st Treatment | 75 | 66 | 87 | 72 | 64 | 84 | 77 | 67 | 87 | 68 | 62 | 78 |
|  | After Last Treatment | 70 | 61 | 79 | 67 | 57 | 74 | 73 | 63 | 80 | 67 | 60 | 77 |
| Bilirubin total (mg/dL) | Hospital Admission | 5,0 | 1,4 | 18,7 | 4,5 | 1,3 | 15,4 | 6,9 | 1,7 | 20,9 | 16,5 | 4,4 | 25,8 |
|  | Baseline | 8,3 | 3,1 | 18,9 | 5,3 | 2,8 | 14,6 | 12,3 | 3,5 | 23,0 | 14,9 | 4,8 | 25,6 |
|  | Post 1st Treatment | 8,5 | 3,5 | 15,9 | 6,3 | 2,9 | 11,7 | 10,8 | 3,6 | 17,9 | 12,6 | 5,1 | 22,2 |
|  | After Last Treatment | 9,4 | 4,1 | 15,7 | 7,5 | 3,8 | 11,6 | 10,6 | 4,3 | 17,7 | 11,8 | 6,1 | 18,9 |
| Potassium (mmol/L) | Hospital Admission | 4,3 | 3,8 | 5,0 | 4,3 | 3,8 | 5,1 | 4,2 | 3,5 | 4,8 | 4,2 | 3,8 | 5,1 |
|  | Baseline | 4,2 | 3,9 | 4,7 | 4,5 | 4,0 | 5,0 | 4,1 | 3,7 | 4,4 | 4,1 | 3,9 | 4,6 |
|  | Post 1st Treatment | 4,3 | 3,9 | 4,5 | 4,3 | 4,0 | 4,7 | 4,2 | 3,9 | 4,4 | 4,3 | 4,0 | 4,5 |
|  | After Last Treatment | 4,4 | 4,1 | 4,8 | 4,6 | 4,1 | 5,0 | 4,3 | 4,0 | 4,7 | 4,4 | 4,1 | 4,9 |
| Sodium (mmol/L) | Hospital Admission | 136 | 131 | 140 | 136 | 131 | 140 | 136 | 131 | 140 | 133 | 127 | 137 |
|  | Baseline | 138 | 135 | 143 | 138 | 135 | 142 | 139 | 135 | 143 | 138 | 133 | 144 |
|  | Post 1st Treatment | 138 | 136 | 141 | 138 | 137 | 141 | 138 | 135 | 141 | 139 | 136 | 141 |
|  | After Last Treatment | 137 | 134 | 141 | 138 | 136 | 142 | 136 | 133 | 140 | 137 | 134 | 142 |
| Creatinine (mg/dL) | Hospital Admission | 1,8 | 1,1 | 3,0 | 2,3 | 1,3 | 3,6 | 1,5 | 0,9 | 2,4 | 2,1 | 1,3 | 4,1 |
|  | Baseline | 1,8 | 1,2 | 2,9 | 2,0 | 1,5 | 3,5 | 1,3 | 0,8 | 2,4 | 1,9 | 1,2 | 3,9 |
|  | Post 1st Treatment | 1,4 | 0,9 | 2,2 | 1,5 | 1,1 | 2,3 | 1,2 | 0,7 | 1,9 | 1,6 | 0,9 | 2,6 |
|  | After Last Treatment | 1,3 | 0,8 | 1,9 | 1,4 | 0,9 | 2,1 | 1,1 | 0,7 | 1,8 | 1,3 | 1,0 | 1,8 |
| BUN (mg/dL) | Hospital Admission | 36 | 20 | 61 | 38 | 24 | 68 | 31 | 16 | 51 | 51 | 25 | 78 |
|  | Baseline | 31 | 18 | 53 | 30 | 18 | 58 | 32 | 17 | 47 | 36 | 19 | 78 |
|  | Post 1st Treatment | 19 | 13 | 34 | 17 | 12 | 31 | 21 | 13 | 34 | 30 | 15 | 45 |
|  | After Last Treatment | 18 | 12 | 31 | 17 | 10 | 28 | 19 | 13 | 31 | 25 | 14 | 36 |
| Chloride (mmol/L) | Hospital Admission | 104 | 97 | 109 | 104 | 97 | 109 | 104 | 99 | 108 | 100 | 96 | 106 |
|  | Baseline | 107 | 103 | 110 | 106 | 102 | 110 | 107 | 104 | 110 | 106 | 102 | 111 |
|  | Post 1st Treatment | 103 | 99 | 107 | 101 | 98 | 105 | 105 | 102 | 108 | 102 | 98 | 107 |
|  | After Last Treatment | 105 | 100 | 107 | 104 | 99 | 106 | 105 | 104 | 107 | 104 | 99 | 106 |
| Erythrocytes / RBC (10<sup>12</sup>/L) | Hospital Admission | 3,2 | 2,7 | 3,9 | 3,1 | 2,7 | 3,7 | 3,2 | 2,8 | 4,0 | 3,1 | 2,5 | 3,6 |
|  | Baseline | 2,7 | 2,4 | 3,2 | 2,8 | 2,4 | 3,3 | 2,6 | 2,4 | 3,1 | 2,5 | 2,2 | 3,1 |
|  | Post 1st Treatment | 2,6 | 2,4 | 3,0 | 2,7 | 2,4 | 3,0 | 2,6 | 2,4 | 2,9 | 2,5 | 2,2 | 2,7 |
|  | After Last Treatment | 2,5 | 2,3 | 2,9 | 2,6 | 2,3 | 2,9 | 2,5 | 2,3 | 2,9 | 2,3 | 2,2 | 2,5 |
| Leucocytes / WBC (10<sup>9</sup>/L) | Hospital Admission | 11,5 | 7,2 | 17,5 | 12,1 | 7,0 | 17,7 | 11,0 | 7,7 | 17,4 | 14,8 | 7,7 | 21,3 |
|  | Baseline | 13,4 | 7,8 | 21,6 | 15,1 | 7,6 | 23,4 | 13,1 | 8,0 | 18,5 | 17,4 | 7,4 | 25,3 |
|  | Post 1st Treatment | 13,7 | 8,4 | 20,1 | 16,4 | 9,6 | 22,0 | 12,9 | 7,6 | 17,8 | 16,1 | 7,5 | 22,0 |
|  | After Last Treatment | 14,0 | 9,0 | 24,0 | 16,0 | 10,0 | 27,0 | 13,0 | 9,0 | 21,0 | 17,0 | 10,0 | 24,0 |
| Platelet Count (10<sup>9</sup>/L) | Hospital Admission | 139 | 71 | 216 | 122 | 65 | 219 | 144 | 86 | 212 | 106 | 52 | 182 |
|  | Baseline | 87 | 44 | 151 | 81 | 46 | 184 | 89 | 39 | 136 | 75 | 42 | 153 |
|  | Post 1st Treatment | 62 | 32 | 117 | 59 | 33 | 129 | 62 | 28 | 108 | 46 | 33 | 100 |
|  | After Last Treatment | 53 | 27 | 100 | 47 | 27 | 94 | 61 | 27 | 111 | 59 | 41 | 94 |
| pH | Hospital Admission | 7,36 | 7,29 | 7,42 | 7,33 | 7,27 | 7,40 | 7,39 | 7,33 | 7,44 | 7,35 | 7,29 | 7,41 |
|  | Baseline | 7,34 | 7,26 | 7,42 | 7,26 | 7,17 | 7,30 | 7,42 | 7,39 | 7,46 | 7,33 | 7,25 | 7,41 |
|  | Post 1st Treatment | 7,41 | 7,35 | 7,46 | 7,39 | 7,31 | 7,45 | 7,43 | 7,38 | 7,48 | 7,41 | 7,38 | 7,46 |
|  | After Last Treatment | 7,40 | 7,33 | 7,44 | 7,38 | 7,27 | 7,43 | 7,41 | 7,37 | 7,45 | 7,40 | 7,27 | 7,44 |
| HCO<sub>3</sub> (serum bicarbonate) (mmol/L) | Hospital Admission | 20,4 | 16,4 | 24,0 | 19,4 | 16,1 | 22,9 | 20,9 | 17,2 | 25,2 | 18,0 | 15,7 | 22,3 |
|  | Baseline | 19,4 | 16,3 | 24,1 | 16,7 | 14,2 | 19,2 | 23,5 | 20,5 | 26,1 | 18,2 | 16,1 | 23,4 |
|  | Post 1st Treatment | 24,6 | 20,9 | 27,6 | 24,0 | 20,0 | 27,8 | 24,8 | 22,1 | 27,2 | 24,0 | 20,9 | 27,3 |
|  | After Last Treatment | 23,5 | 19,8 | 27,0 | 23,2 | 16,9 | 27,3 | 23,6 | 21,0 | 26,8 | 23,5 | 18,2 | 26,9 |
| pCO<sub>2</sub> (mmHg) | Hospital Admission | 36 | 31 | 42 | 38 | 31 | 43 | 34 | 29 | 41 | 33 | 26 | 41 |
|  | Baseline | 37 | 31 | 43 | 39 | 32 | 46 | 35 | 31 | 41 | 35 | 29 | 44 |
|  | Post 1st Treatment | 39 | 33 | 44 | 40 | 35 | 47 | 36 | 32 | 43 | 36 | 31 | 45 |
|  | After Last Treatment | 37 | 34 | 44 | 39 | 34 | 47 | 37 | 34 | 42 | 37 | 33 | 43 |
| Base Excess (mmol/L) | Hospital Admission | -5,0 | -9,8 | -0,3 | -5,9 | -10,8 | -1,3 | -4,2 | -8,6 | 1,1 | -7,8 | -12,7 | -2,7 |
|  | Baseline | -5,6 | -10,5 | -0,4 | -10,3 | -13,8 | -6,7 | -0,8 | -4,8 | 2,1 | -7,6 | -11,2 | -1,4 |
|  | Post 1st Treatment | 0,2 | -4,0 | 3,5 | -0,5 | -5,1 | 4,0 | 0,6 | -2,8 | 3,4 | 0,1 | -4,4 | 3,2 |
|  | After Last Treatment | -1,0 | -6,0 | 3,3 | -1,2 | -10,1 | 3,8 | -0,8 | -3,5 | 3,0 | -1,2 | -8,4 | 3,2 |
| Noradrenaline highest dose (µg/kg/min) | Hospital Admission | 0,359 | 0,122 | 0,840 | 0,518 | 0,147 | 0,985 | 0,289 | 0,101 | 0,638 | 0,381 | 0,079 | 0,800 |
|  | Baseline | 0,336 | 0,137 | 0,802 | 0,566 | 0,229 | 1,025 | 0,195 | 0,051 | 0,365 | 0,419 | 0,170 | 0,914 |
|  | Post 1st Treatment | 0,333 | 0,110 | 0,837 | 0,497 | 0,181 | 0,918 | 0,180 | 0,059 | 0,453 | 0,350 | 0,100 | 0,800 |
|  | After Last Treatment | 0,207 | 0,083 | 0,511 | 0,244 | 0,108 | 0,528 | 0,174 | 0,040 | 0,495 | 0,195 | 0,089 | 0,453 |
| Albumin (g/L) | Hospital Admission | 22 | 17 | 27 | 21 | 16 | 25 | 23 | 18 | 27 | 20 | 17 | 25 |
|  | Baseline | 22 | 17 | 27 | 21 | 16 | 26 | 23 | 17 | 27 | 23 | 17 | 27 |
|  | Post 1st Treatment | 22 | 18 | 28 | 22 | 17 | 27 | 24 | 18 | 29 | 22 | 18 | 28 |
|  | After Last Treatment | 20 | 14 | 26 | 19 | 13 | 25 | 22 | 15 | 27 | 19 | 16 | 25 |
| Lactate (mmol/L) | Hospital Admission | 3,3 | 2,0 | 7,1 | 3,6 | 2,2 | 7,3 | 2,7 | 1,9 | 6,9 | 3,6 | 2,1 | 7,0 |
|  | Baseline | 4,4 | 2,0 | 10,0 | 7,9 | 3,0 | 12,4 | 2,4 | 1,6 | 5,2 | 4,4 | 2,0 | 8,6 |
|  | Post 1st Treatment | 3,3 | 1,9 | 9,9 | 6,2 | 2,1 | 13,6 | 2,5 | 1,5 | 3,8 | 3,4 | 2,1 | 7,2 |
|  | After Last Treatment | 3,2 | 1,8 | 11,6 | 5,2 | 2,1 | 15,5 | 2,0 | 1,6 | 6,2 | 5,7 | 1,7 | 15,8 |
| Acidosis (n, % < pH 7.35) | Hospital Admission | 109 | 39% |  | 77 | 53% |  | 32 | 24% |  | 22 | 42% |  |
|  | Baseline | 146 | 52% |  | 146 | 100% |  | 0 | 0% |  | 31 | 58% |  |
|  | Post 1st Treatment | 58 | 21% |  | 40 | 27% |  | 18 | 13% |  | 10 | 19% |  |
|  | After Last treatment | 43 | 15% |  | 48 | 33% |  | 12 | 9% |  | 11 | 21% |  |

**Table 6.**
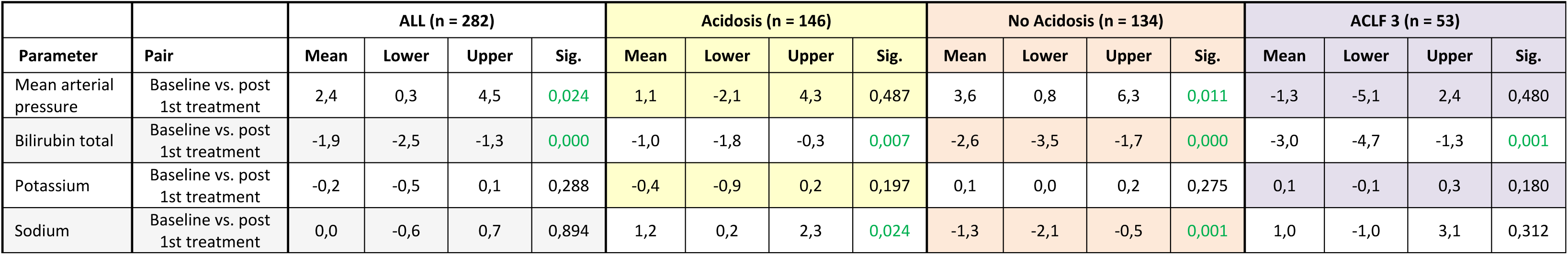
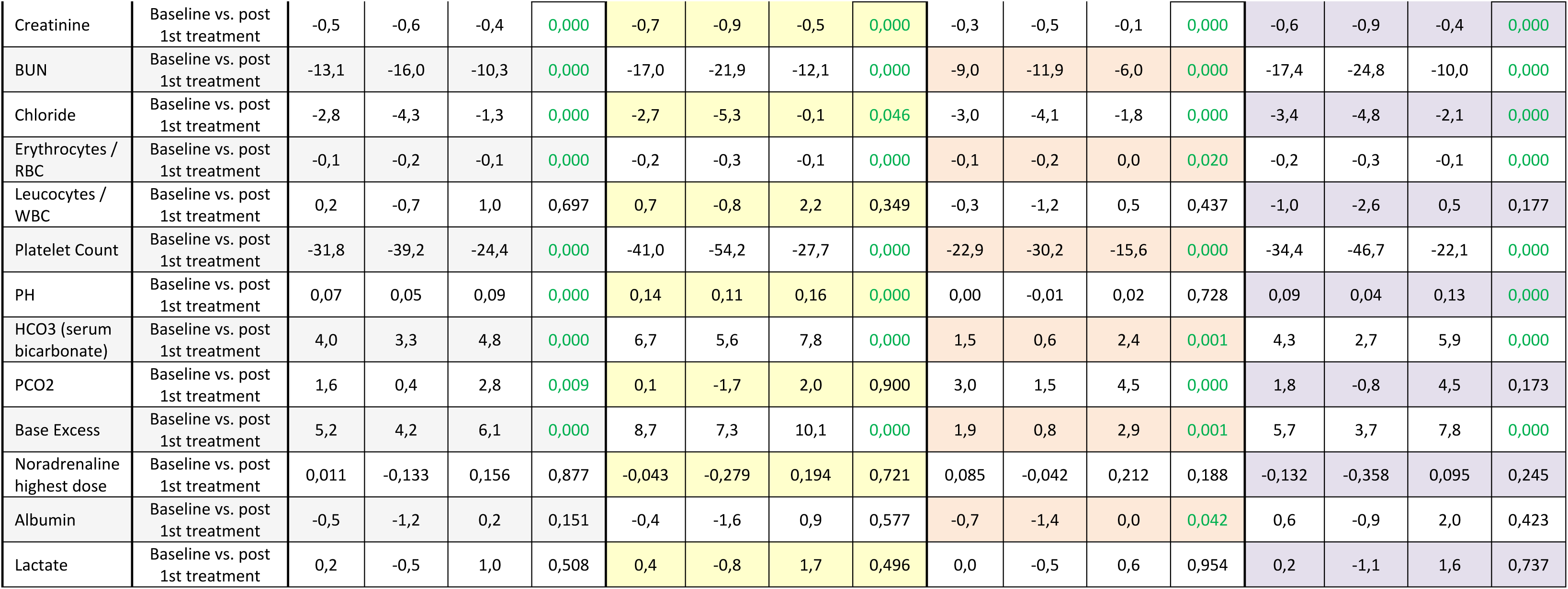
Paired differences of performance parameters between baseline and after the first treatment in the whole data set and in each of the subgroups. Patients dead without data recording after the first ADVOS treatment session are excluded from the analysis since no pairing was possible.s

| Parameter | Pair | ALL (n = 282) |  |  |  | Acidosis (n = 146) |  |  |  | No Acidosis (n = 134) |  |  |  | ACLF 3 (n = 53) |  |  |  |
| --- | --- | --- | --- | --- | --- | --- | --- | --- | --- | --- | --- | --- | --- | --- | --- | --- | --- |
|  |  | Mean | Lower | Upper | Sig. | Mean | Lower | Upper | Sig. | Mean | Lower | Upper | Sig. | Mean | Lower | Upper | Sig. |
| Mean arterial pressure | Baseline vs. post 1st treatment | 2,4 | 0,3 | 4,5 | 0,024 | 1,1 | -2,1 | 4,3 | 0,487 | 3,6 | 0,8 | 6,3 | 0,011 | -1,3 | -5,1 | 2,4 | 0,480 |
| Bilirubin total | Baseline vs. post 1st treatment | -1,9 | -2,5 | -1,3 | 0,000 | -1,0 | -1,8 | -0,3 | 0,007 | -2,6 | -3,5 | -1,7 | 0,000 | -3,0 | -4,7 | -1,3 | 0,001 |
| Potassium | Baseline vs. post 1st treatment | -0,2 | -0,5 | 0,1 | 0,288 | -0,4 | -0,9 | 0,2 | 0,197 | 0,1 | 0,0 | 0,2 | 0,275 | 0,1 | -0,1 | 0,3 | 0,180 |
| Sodium | Baseline vs. post 1st treatment | 0,0 | -0,6 | 0,7 | 0,894 | 1,2 | 0,2 | 2,3 | 0,024 | -1,3 | -2,1 | -0,5 | 0,001 | 1,0 | -1,0 | 3,1 | 0,312 |
| Creatinine | Baseline vs. post 1st treatment | -0,5 | -0,6 | -0,4 | 0,000 | -0,7 | -0,9 | -0,5 | 0,000 | -0,3 | -0,5 | -0,1 | 0,000 | -0,6 | -0,9 | -0,4 | 0,000 |
| BUN | Baseline vs. post 1st treatment | -13,1 | -16,0 | -10,3 | 0,000 | -17,0 | -21,9 | -12,1 | 0,000 | -9,0 | -11,9 | -6,0 | 0,000 | -17,4 | -24,8 | -10,0 | 0,000 |
| Chloride | Baseline vs. post 1st treatment | -2,8 | -4,3 | -1,3 | 0,000 | -2,7 | -5,3 | -0,1 | 0,046 | -3,0 | -4,1 | -1,8 | 0,000 | -3,4 | -4,8 | -2,1 | 0,000 |
| Erythrocytes / RBC | Baseline vs. post 1st treatment | -0,1 | -0,2 | -0,1 | 0,000 | -0,2 | -0,3 | -0,1 | 0,000 | -0,1 | -0,2 | 0,0 | 0,020 | -0,2 | -0,3 | -0,1 | 0,000 |
| Leucocytes / WBC | Baseline vs. post 1st treatment | 0,2 | -0,7 | 1,0 | 0,697 | 0,7 | -0,8 | 2,2 | 0,349 | -0,3 | -1,2 | 0,5 | 0,437 | -1,0 | -2,6 | 0,5 | 0,177 |
| Platelet Count | Baseline vs. post 1st treatment | -31,8 | -39,2 | -24,4 | 0,000 | -41,0 | -54,2 | -27,7 | 0,000 | -22,9 | -30,2 | -15,6 | 0,000 | -34,4 | -46,7 | -22,1 | 0,000 |
| PH | Baseline vs. post 1st treatment | 0,07 | 0,05 | 0,09 | 0,000 | 0,14 | 0,11 | 0,16 | 0,000 | 0,00 | -0,01 | 0,02 | 0,728 | 0,09 | 0,04 | 0,13 | 0,000 |
| HCO3 (serum bicarbonate) | Baseline vs. post 1st treatment | 4,0 | 3,3 | 4,8 | 0,000 | 6,7 | 5,6 | 7,8 | 0,000 | 1,5 | 0,6 | 2,4 | 0,001 | 4,3 | 2,7 | 5,9 | 0,000 |
| PCO2 | Baseline vs. post 1st treatment | 1,6 | 0,4 | 2,8 | 0,009 | 0,1 | -1,7 | 2,0 | 0,900 | 3,0 | 1,5 | 4,5 | 0,000 | 1,8 | -0,8 | 4,5 | 0,173 |
| Base Excess | Baseline vs. post 1st treatment | 5,2 | 4,2 | 6,1 | 0,000 | 8,7 | 7,3 | 10,1 | 0,000 | 1,9 | 0,8 | 2,9 | 0,001 | 5,7 | 3,7 | 7,8 | 0,000 |
| Noradrenaline highest dose | Baseline vs. post 1st treatment | 0,011 | -0,133 | 0,156 | 0,877 | -0,043 | -0,279 | 0,194 | 0,721 | 0,085 | -0,042 | 0,212 | 0,188 | -0,132 | -0,358 | 0,095 | 0,245 |
| Albumin | Baseline vs. post 1st treatment | -0,5 | -1,2 | 0,2 | 0,151 | -0,4 | -1,6 | 0,9 | 0,577 | -0,7 | -1,4 | 0,0 | 0,042 | 0,6 | -0,9 | 2,0 | 0,423 |
| Lactate | Baseline vs. post 1st treatment | 0,2 | -0,5 | 1,0 | 0,508 | 0,4 | -0,8 | 1,7 | 0,496 | 0,0 | -0,5 | 0,6 | 0,954 | 0,2 | -1,1 | 1,6 | 0,737 |

#### Normalization and improvement of the blood composition in case of electrolyte disturbances or acid-base disorders

All the electrolytes remained within the expected physiological range during treatments (Table 5).

Acid-base parameters significantly improved. pH (7.34 vs. 7.41, p<0.001), HCO_3_ ^-^ (19.4 vs. 24.6 mmol/l, p<0.001) and base excess (–5.6 vs. 0.2 mmol/l, p<0.001) returned to the physiological range after the first ADVOS session (Table 5). In the subgroup of patients with acidosis at baseline, an even higher correction of acid-base balance was achieved for pH (7.26 vs. 7.39, p<0.001), serum bicarbonate (16.7 vs. 24.0 mmol/l, p<0.001) and base excess (–10.3 vs. –0.5 mmol/l, p<0.001). Moreover, only 27% of the patients with acidosis at baseline had a pH < 7.35 after the first ADVOS session (146 vs. 40 patients) (Table 5).

Results from the subgroup of patients with respiratory acidosis are shown in Supplementary Table 5 and 6. The CO2 removal rate could not be calculated in these patients due to the absence of routine post-dialyzer sampling.

#### Removal of fluid

An average ultrafiltration rate and volume of 135 (IQR 70, 255) mL and 2500 (IQR 650, 4882) mL were achieved, respectively (Table 2).

### 3.4 Mortality rate and SOFA Standardized Mortality Ratio

The documented mortality rate among the 282 patients in the ADVOS registry reached 59.6% and 67.7% 28 and 90 days after the first ADVOS treatment session, respectively.

202 patients had a documented SOFA Score at baseline and were eligible for the calculation of the SMR. The median SOFA Score is these patients was 15 (IQR 12, 18) and 91% had multiple organ failure. According to the SOFA Score 169 deaths (84%) were expected, while 134 (66%) were observed among the 202 patients with an available SOFA Score (Figure 1). This translates into a SMR of 0.79 (CI 95%: 0.66-0.93) with a number needed to treat (NNT) of 5.8. The absolute risk reduction was 17% and was higher in the subgroup of patients with SOFA Scores between 12 and 14 (AAR of 29%, data not shown). Patients with acidosis showed SMR, AAR and NNT of 0.90 (CI95%: 0.70-1.10), 9% and 11.7, respectively. In the case of ACLF 3, similar values to the overall population were observed with a SMR of 0.80 (CI95%: 0.53-1.06), an AAR of 18% and a NNT of 4.5 (Supplementary Table 3).

**Figure 1.**
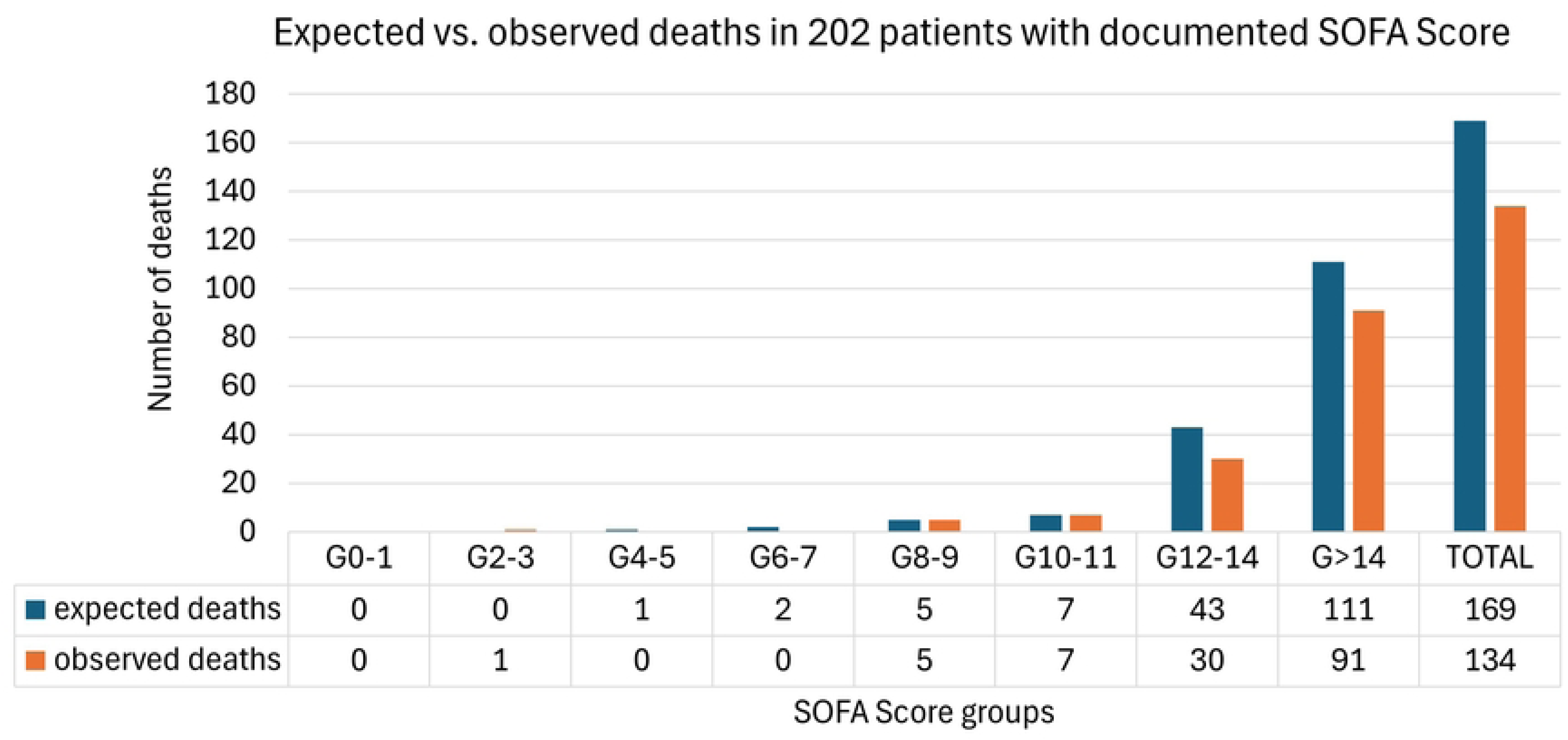
Expected deaths according to SOFA Score immediately before the first ADVOS session (blue) vs. observed deaths at the end of the registry (red) among different subgroups of SOFA score based on the publication from Ferreira et al [23].

## 4. Discussion

### Key results

The current analysis demonstrates the feasibility and safety of the ADVOS hemodialysis system in the larger cohort to date. The intended purpose to remove water-soluble and protein-bound substances, normalize blood composition (including acidosis correction) and remove fluid was fulfilled in the intended population of 282 patients with acute, chronic and acute-on-chronic liver failure and/or renal failure and/or acidosis. Patients were treated under real-life conditions without any specific study-related intervention. This resulted in an adequate safety profile with a low rate of adverse events directly related to the ADVOS hemodialysis system.

ADVOS can simultaneously support three organs (i.e., liver, kidney, and lung) and concurrently correct acid-base imbalances. In this report, a reduction in renal clinical markers such as creatinine (up to 26%) or BUN (up to 33%) has been shown. These data are within expected ranges reported in larger randomized control trials with benchmark hemodialysis devices for patients with acute kidney injury. In these studies, depending on the modality employed (intermittent or continuous), the intensity (higher vs. lower doses) or the timing (early vs. delayed), the reduction of creatinine and BUN ranged from 9 to 81% and 17 to 71%, respectively [28–31]. In the case of bilirubin, a concentration dependent reduction had already been shown with ADVOS [7]. In our analysis, bilirubin levels were reduced up to 20% in patients with ACLF grade 3 after one single ADVOS session. The reference study by Bañares et al. with the MARS extracorporeal liver support system achieved a reduction of 26% with higher median starting bilirubin levels [32]. In each of the reported case series with ADVOS, similar levels were also reached [5, 33, 34].

Acid-base correction has also been achieved as shown by the changes in blood gas values. This was especially meaningful in the subgroup of patients with acidosis, where only 27% had still a pH < 7.35 after the first ADVOS session. To note, at the time patients were treated along the EMOS Registry, only concentrates with full bicarbonate content (i.e., BIC 20) were available, which allow a lower bicarbonate gradient between blood and dialysate and probably a lower performance in terms of CO_2_ removal as BIC 10 or BIC 0. Therefore, acidosis correction was mainly driven by changes in bicarbonate, except for the subgroup of patients with respiratory acidosis (Supplementary Tables 5 and 6). Previously published case series in patients with multiorgan failure or with COVID-19 treated with ADVOS already reported improvement of the acid-base balance. Indeed, a correction of acidosis within 6 hours and a median CO_2_ removal of 49 mL/min was documented, respectively [7, 10]. Very few randomized controlled trials have published similar data. In a subgroup analysis of the RENAL study, 50% of patients had no acidosis after 24 hours [35]. In larger studies using sodium bicarbonate or extracorporeal CO_2_ removal (ECCO2R) this percentage of patients with corrected acidosis was either not documented or not reached [36, 37].

Additionally, the evaluation of the standardized mortality ratio serves as a quality marker in registry data and as quality measure in health care systems. In our current analysis a significant difference was observed between the expected mortality rate according to the SOFA score and the observed mortality. In 202 patients with a documented SOFA Score before the first ADVOS treatment a SMR of 0.79 (CI 95%: 0.66-0.93) with an absolute risk reduction of 17% was observed. This resulted in a NNT of 5.8, which seems to improve numbers documented in a meta-analysis for other similar devices [38]. Although several drawbacks have been reported [39], risk-adjusted mortality analyses are commonly used to assess and compare treatment quality [40, 41]. In the current registry, the stratification based on the highest SOFA proposed by Ferreira et al. score was employed [23]. In the latter as well as in a systematic review published afterwards, studies that evaluated the prognostic value of highest SOFA scores during ICU stay found excellent discrimination [20].

### Interpretation and generalizability

The ADVOS hemodialysis system has been available in the German market since 2013. According to data provided by the manufacturer, in the last decade more than 6000 treatment sessions in more than 20 different hospitals have been performed. Considering the median treatment sessions per patient documented in this analysis, we estimate that around 2000 patients could have already been treated with ADVOS. In this period, no relevant safety issues have been identified. Moreover, the published literature shows the feasibility of the therapy and the fulfillment of the intended purpose [2].

The current analysis of the EMOS-Registry accounts for approximately 15% of the treated patients. This results in a highly valuable tool for the interpretation of real-world data of the ADVOS therapy. It is hence expected that these results could be generalizable. However, due to the broad spectrum of patient groups that could potentially benefit from ADVOS, a better understanding of each of the specific population is needed. We depicted data from patients with acidosis and ACLF grade 3 and found several differences in terms of therapy settings (i.e., dialysate pH, 7.4 vs. 8.0), performance (i.e., bilirubin reduction ratio, 14% vs. 3%), or need for mechanical ventilation at baseline (58% vs, 76%), among others. This indicates a need for adapted individualized treatment in each of the subgroups. Moreover, the median SOFA Score of 15 indicates that ADVOS might have been used as a rescue therapy, probably due to the novelty of the therapy at the time of conducting the registry. Therefore, present data needs to be endorsed by randomized controlled trials. In this regard, several prospective studies were being performed at the time of writing this report, including studies in patients with acidosis (NCT05842369), with high vasopressor demand (DRKS00031279) or with liver dysfunction (NCT06129617).

### Limitations

As expected in this kind of studies, due to its retrospective nature, some inherent level of selection bias cannot be ruled out [42]. However, the result of this analysis agrees with previously reported data from other cohorts [5, 7, 8, 10, 34, 43], which gives validity to documented outcomes.

External validity might also be impaired since all data was obtained from patients from a single country. Moreover, the different contributions of each of the participating centers, as well as the different type of department where the treatments were performed (ICU vs. dialysis unit) could have impacted the outcome. In any case, all the data presented regarding the feasibility and safety of the ADVOS hemodialysis device were observed separately in each of the clinical sites too (data not shown).

A re-evaluation of patients terminating the registry alive before day 90 and not being in hospital at that point was not possible in the absence of informed consent. These patients had been discharged alive from ICU and/or hospital. Thus, they were considered as still alive on day 90 for calculation purposes. This could have influenced the accurate calculation of the SMR. Moreover, the SMR based on SOFA Score refers to data published in 2001, and predicted mortality might be currently different. However, until a better alternative is proposed, the SOFA Score is still commonly accepted.

Many of the patients included in this analysis were treated during the first years after the introduction of the ADVOS therapy in the hospitals. Therefore, the current expertise and internal guidelines available in each of the centers now were probably not yet properly implemented. It is expected that with the acquired knowledge the selection of patients and the application of the ADVOS hemodialysis system should be more adequate to obtain better clinical results.

## 5. Conclusion

The final report of the Registry on Extracorporeal Multiple Organ Support (EMOS) with the ADVOS hemodialysis system demonstrated the fulfillment of the intended purpose to remove water-soluble and protein-bound substances and to correct acid-base balance in a large heterogeneous cohort of 282 patients with multiorgan failure, including those with acute-on-chronic liver failure, acidosis and an indication for dialysis. Additionally, a significantly reduced SOFA Score-standardized mortality ratio was documented. These data need to be confirmed in prospective randomized controlled trials covering each of the subgroups that could potentially benefit from the ADVOS therapy.

## 7. Declarations

### 7.1 Ethics approval and consent to participate

All patients who participated in the registry and for whom pseudonymous data collection was scheduled signed an informed consent form. An authorized investigator who had been trained on the registry explained to the patients the content and meaning of the registry. For patients who were unable or unwilling to provide informed consent and for patients whose data had been collected retrospectively, data was anonymous (i.e., untraceable) and could not be allocated to a patient’s name. The registry was first approved by the Ethics Committee of the Bavarian Medical Association (BLÄK: Bayerische Landesärztekammer) in their meeting Nr. 16022 on the 12^th^ of May 2016. Approval of Regional or local Ethics Committees for subsequently participating centers was obtained.

### 7.2 Consent for publication

Patients whose data was pseudonymously recorded signed an informed consent form. Consent was waived for patients whose data was anonymously recorded.

### 7.3 Availability of data and materials

The datasets used and/or analyzed during the current study are available from the corresponding author on reasonable request.

### 7.4 Competing interests

JWM and VF received research funding from ADVITOS GmbH. AP and TB are employed by ADVITOS GmbH. Other authors declare that they have no conflict of interests.

### 7.5 Funding

This project has received funding from the European Union’s Horizon 2020 research and innovation programme under grant agreement No 880349.

### 7.6 Authors’ contributions

VF and JL participated in the study design and conceptualization. AP, TMB, BT and VF contributed with the statistical analysis and wrote the manuscript. AF, DJ, JL, BT, PK, KC, OB and MS participated in data curation. JWM, AK and SK provided critical review with important intellectual input. All authors are accountable for all aspects of the work, and all authors read and approved the final manuscript.

## 6. List of abbreviations

AE: adverse event
ADVOS: Advanced Organ Support System
ARR: absolute risk reduction
BUN: blood urea nitrogen
CO_2_: Carbon dioxide
DRKS: German Registry for Clinical Studies
GCS: Glasgow coma score
HCO^-^: bicarbonate
ICU: intensive care unit
MOF: multiple organ failure
NNT: number needed to treat
pCO_2_: partial pressure of carbon dioxide
SOFA: sequential organ failure assessment
SMR: standardized mortality ratio
UF: Ultrafiltration

## 7.7 Acknowledgements

In memoriam of Bernhard Kreymann and Wolfgang Huber in appreciation of their invaluable contribution to both the development of ADVOS therapy and its implementation, as well as their guidance in the design of the present study.

## Notes

### Competing Interest Statement

JWM and VF received research funding from ADVITOS GmbH. AP and TMB are employed by ADVITOS GmbH. Other authors declare that they have no conflict of interests.

### Clinical Trial

DRKS00017068

### Funding Statement

Yes

### Author Declarations

All patients who participated in the registry and for whom pseudonymous data collection was scheduled signed an informed consent form. An authorized investigator who had been trained on the registry explained to the patients the content and meaning of the registry. For patients who were unable or unwilling to provide informed consent and for patients whose data had been collected retrospectively, data was anonymous (i.e., untraceable) and could not be allocated to a patient’s name. The registry was first approved by the Ethics Committee of the Bavarian Medical Association (BLÄK: Bayerische Landesärztekammer) in their meeting Nr. 16022 on the 12th of May 2016. Approval of Regional or local Ethics Committees for subsequently participating centers was obtained.

